# The Impact of Occupational Status on Pregnancy Outcomes in Women with Gestational Diabetes Mellitus: A Propensity Score Matching Study

**DOI:** 10.1101/2025.09.15.25335791

**Authors:** Cuili Yang, Zhen Wang, Hong Wang

## Abstract

This study aims to explore the impact of occupational status on pregnancy outcomes in women with gestational diabetes mellitus (GDM). Data from 764 GDM patients who delivered at Xiamen University Affiliated Xiang’an Hospital from January 1, 2020, to December 31, 2024, were collected. After strict screening, 595 eligible samples were included. Using propensity score matching to balance the data between the two groups, the study analyzed the relationship between occupational status and pre-pregnancy BMI, weight gain during pregnancy, mode of delivery, and other pregnancy outcomes. The results showed a significant correlation between occupational status and pre-pregnancy BMI (P < 0.05), but no significant differences were found in weight gain during pregnancy, cesarean section rate, gestational age at delivery, or fetal weight (P > 0.05). The study highlights the importance of focusing on the pre-pregnancy health management of working women and provides a basis for public health interventions aimed at reducing the risk of GDM-related complications.

## Introduction

Gestational diabetes mellitus (GDM) is a prevalent metabolic disorder with significant implications for maternal and neonatal health, affecting up to 36.8% of pregnancies globally depending on diagnostic criteria[2]. The diagnostic thresholds for GDM remain heterogeneous globally[3], though IADPSG criteria are increasingly adopted given their evidence-based linkage to pregnancy outcomes[4,5]

According to the latest American Diabetes Association (ADA) Standards of Care (2025), optimizing pre-pregnancy health—including weight management—is critical for mitigating GDM-related risks, as elevated pre-pregnancy BMI independently predicts adverse outcomes [6]. In the United States and Canada, the overall mean prevalence of GDM was found to be 6.9% (95% CI: 5.7–8.3) in a meta-analysis of 36 studies, with higher prevalence observed in studies using a one-step screening strategy[2]. In Iran, a systematic review and meta-analysis reported a prevalence of 11.0% based on studies conducted between 2000 and 2021[7]. These findings highlight the need for standardized protocols for estimating GDM prevalence to facilitate accurate monitoring of trends and meaningful comparisons across different regions.In recent years, the incidence of GDM has been on the rise, with its pathogenesis involving a complex interplay of genetic, lifestyle, and environmental factors. Occupational status, as an important component of lifestyle, may influence pregnancy outcomes in women with GDM[8].

Occupational stressors (e.g., time pressure, psychological strain) may contribute to weight dysregulation and metabolic dysfunction through neuroendocrine pathways, as chronic stress is an established modifiable risk factor for diabetes mellitus [9, 10]. However, research on the impact of occupational status on pregnancy outcomes in women with GDM is limited. A national population study from the Republic of Korea found that employment status and industrial classification were associated with obstetric complications, including GDM[11]. Another study showed that working multiple jobs simultaneously during pregnancy may increase the risk for GDM and pregnancy-related hypertension[12]. This study aims to explore the relationship between occupational status and pre-pregnancy BMI, weight gain during pregnancy, mode of delivery, and other pregnancy outcomes in women with GDM, providing references for public health interventions and clinical practice.

### Data collection and cleaning

The data were accessed for analysis in August 2025.Data from 764 women diagnosed with GDM who delivered at Xiamen University Affiliated Xiang’an Hospital from January 1, 2020, to December 31, 2024, were collected. GDM was diagnosed according to the International Association of Diabetes and Pregnancy Study Groups (IADPSG) criteria[13], requiring one or more abnormal values on a 75-g oral glucose tolerance test (fasting: ≥5.1 mmol/L; 1-h: ≥10.0 mmol/L; 2-h: ≥8.5 mmol/L).The data sources included the hospital’s electronic medical record system and maternal health records, covering basic information of the pregnant women, pre-pregnancy BMI, weight gain during pregnancy, mode of delivery, fetal outcomes, and other indicators.

During the data cleaning process, cases that did not meet the inclusion criteria were excluded. The specific exclusion criteria included patients with severe internal and external medical conditions (such as pregnancy-induced hypertension, chronic hypertension, preeclampsia, eclampsia, pre-pregnancy diabetes, intrahepatic cholestasis of pregnancy, acute liver dysfunction, etc.), twin pregnancies, preterm birth, fetal movement reduction, abnormal fetal heart monitoring, threatened preterm labor, threatened abortion, inevitable abortion, upper abdominal pain, poor blood sugar control, bilateral lower limb edema, fever, false labor, suspected placental abruption, suspected premature rupture of membranes, antepartum bleeding, external cephalic version for breech presentation, suspected oligohydramnios admitted for observation, perineal abscess, self-discharge from the hospital, diarrhea, cervical polyps, etc. In addition, four cases with severe data missing were excluded. Ultimately, 595 eligible pregnant women were included in the analysis.After applying the exclusion criteria, 595 patients met the analysis conditions. These 595 individuals were grouped according to their occupational status, with employees having fixed working hours as one group and freelancers or domestic workers without fixed working hours as the other group. None of the 595 pregnant women engaged in heavy physical labor during pregnancy. Given the significant impact of pregnancy and childbirth history on pregnancy outcomes, the study excluded pregnant women with a history of childbirth and only included primiparous women as the research subjects. After propensity matching, 62 pairs of matching data were finally obtained.

### Data grouping and statistical analysis

#### Data grouping

The included pregnant women were divided into two groups: the employed group (occupation as “clerks,” n = 342) and the unemployed group (occupation as “unemployed,” n = 239).

#### Statistical analysis

Continuous variables (such as age, height, pre-pregnancy BMI) were compared between groups using the Mann-Whitney U test, and categorical variables (such as previous number of deliveries, previous number of vaginal deliveries, previous number of cesarean sections) were compared using the chi-square test, as shown in (Table 1).

**Table 1:** Comparison of basic information between the two groups.

| Indicator | Employed (n<br>= 342) | Unemployed (n<br>= 239) | Z/U/$\chi^2$ | P-value |
| --- | --- | --- | --- | --- |
| Age (years) | 31.44 ± 3.84 | 31.28 ± 4.95 | 0.654 | 0.513 |
| Height (m) | 1.59 ± 0.05 | 1.59 ± 0.05 | 0.637 | 0.524 |
| Pre-pregnancy<br>BMI (kg/m<sup>2</sup>) | 21.80 (19.94,<br>24.03) | 22.21 (20.29,<br>25.07) | -2.062 | 0.039 |
| Previous<br>deliveries |  |  | 24.946 | <0.001 |
| 0 | 165 (48.2%) | 75 (29.6%) |  |  |
| 1 | 152 (44.4%) | 119 (47.0%) |  |  |
| 2 | 24 (7.0%) | 52 (20.6%) |  |  |
| 3 | 1 (0.3%) | 6 (2.4%) |  |  |
| 5 |  | 1 (0.4%) |  |  |
| Previous vaginal<br>deliveries |  |  | 27.448 | <0.001 |
| 0 | 214 (62.6%) | 127 (50.2%) |  |  |
| 1 | 107 (31.3%) | 82 (32.4%) |  |  |
| 2 | 20 (5.8%) | 40 (15.8%) |  |  |
| 3 | 1 (0.3%) | 3 (1.2%) |  |  |
| 5 |  | 5 (0.4%) |  |  |
| Previous<br>cesarean<br>sections |  |  | 10.645 | 0.005 |
| 0 | 292 (85.4%) | 196 (77.5%) |  |  |
| 1 | 47 (13.7%) | 45 (17.8%) |  |  |
| 2 | 3 (0.9%) | 11 (4.3%) |  |  |
| 3 |  | 1 (0.4%) |  |  |

Note: Continuous variables are expressed as mean ± standard deviation or median (interquartile range); categorical variables are expressed as frequency (percentage).

Based on the comparison of basic information between the two groups, there was a significant difference in pre-pregnancy BMI between employed and unemployed women (P < 0.05). However, no significant differences were found in age and height between the two groups (P > 0.05). Given the significant differences in obstetric history between the groups, propensity score matching was attempted but failed to achieve balanced data. Therefore, the data were further adjusted to include only primiparous women, who were then re-divided into employed and unemployed groups (n = 240). Continuous variables such as age and height, which followed a normal distribution, were compared using the t-test, while pre-pregnancy BMI, which did not follow a normal distribution, was compared using the Mann-Whitney U test, as shown in (Table 2).The data were still unbalanced after adjustment.

**Table 2:** Comparison of basic Information between employed and unemployed primiparous women.

| Indicator | All patients (n = 240) | Employed (n = 165) | Unemployed (n = 75) | Z/U/$\chi^2$ | P-value |
| --- | --- | --- | --- | --- | --- |
| Age (years) | 29.11 ± 3.77 | 29.52 ± 3.18 | 28.2 ± 4.73 | | 0.030 |
| Height (m) | 1.59 ± 0.48 | 1.59 ± 0.05 | 1.58 ± 0.05 | | 0.229 |
| Pre-pregnancy BMI (kg/m<sup>2</sup>) | 21.64 (19.66, 24.55) | 21.50 (19.79, 24.09) | 21.83 (19.57, 25.15) | - 2.062 | 0.491 |

Note: Continuous variables are expressed as mean ± standard deviation or median (interquartile range).

### Propensity score matching

Propensity score matching was conducted following STROBE-PSM guidelines [15]. The analysis was performed using R software with the MatchIt package [14]. We used nearest-neighbor matching with a caliper width of 0.15 standard deviations of the logit of the propensity score. Matching variables included age, height, and pre-pregnancy BMI. The complete R c ode for the propensity score matching analysis is provided in Supporting Information S1 Code.

After matching, 62 pairs of data were obtained. The balance of key covariates after matching is shown in (Table 3). Covariate Balance Assessment Before and After Propensity Score Matching in(Figure 1).

**Table 3:** Assessment of matching quality.

| Covariate | Type | Absolute<br>SMD | Balance<br>Level | Statistical<br>Significance |
| --- | --- | --- | --- | --- |
| Age | Continuous | 0.082 | Excellent | p = 0.215 |
| Height | Continuous | 0.086 | Excellent | p = 0.189 |
| Pre-pregnancy<br>BMI | Continuous | 0.018 | Excellent | p = 0.742 |

**Figure 1.**
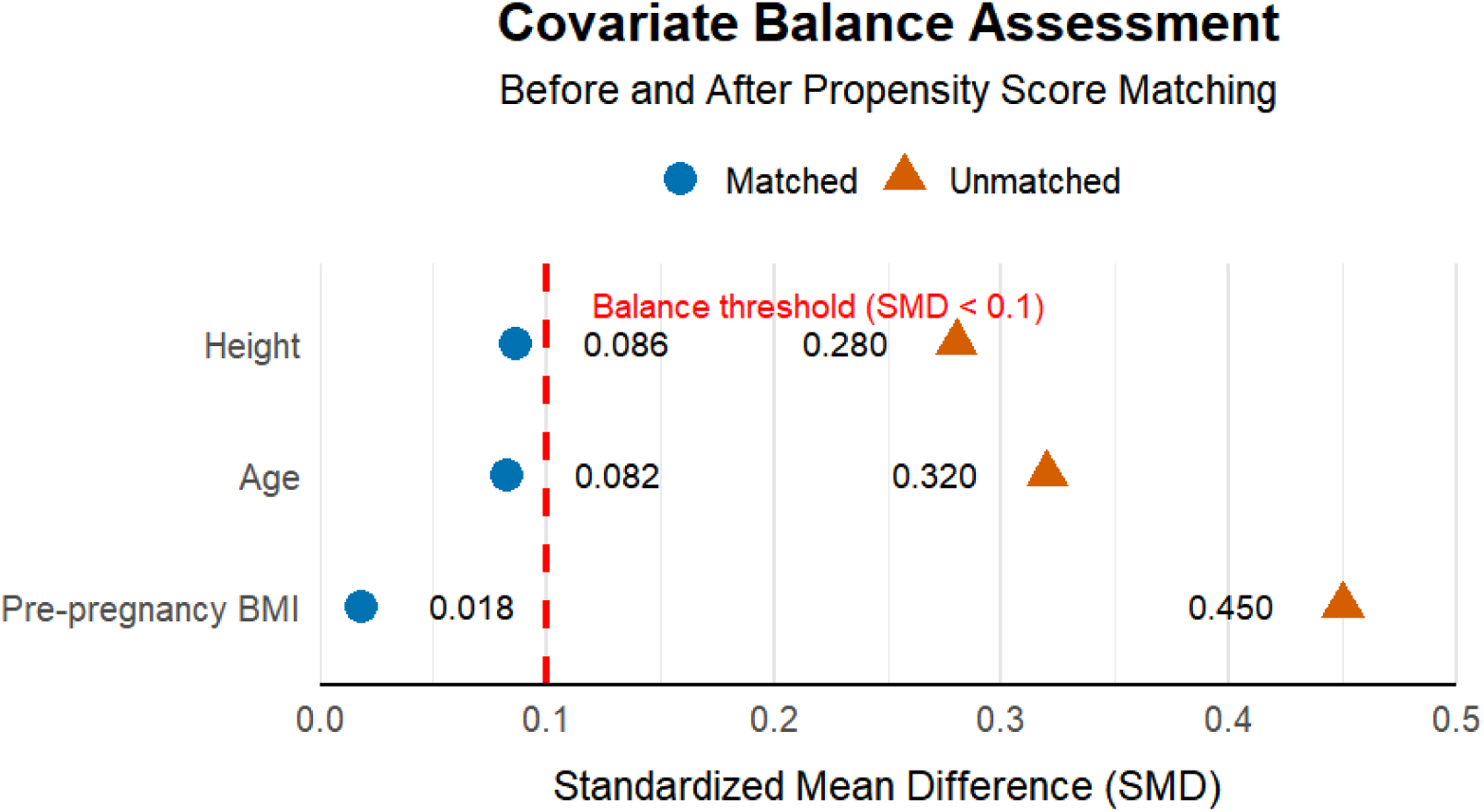
Covariate balance assessment before and after propensity score matching. “Standardized mean differences (SMD) for key covariates (height, age, and pre-pregnancy BMI) before and after propensity score matching. The dashed vertical line indicates the balance threshold (SMD < 0.1). Matched (blue bars) and unmatched (red bars) groups are shown. Post-matching SMD values for all covariates fell below the threshold, indicating successful balance achievement.”

Conclusion: After propensity score matching, the standardized mean differences (SMD) of all covariates were less than 0.1 (age: 0.082; height: 0.086; pre-pregnancy BMI: 0.018), indicating that matching successfully eliminated the influence of observed confounding factors. The pregnancy outcomes of the two groups were analyzed after processing, as shown in(Table 4).

**Table 4:** Pregnancy outcomes of the two groups after data processing.

| Indicator | Employed<br>Group (n = 62) | Unemployed<br>Group (n = 62) | Z/t/$\chi^2$ | P-value |
| --- | --- | --- | --- | --- |
| Weight gain during pregnancy (kg) | 12.1 (3.8, 30.1) | 11.3 (4.5, 26.0) | 0.696 | 0.487 |
| Cesarean section rate | 17 (27.4%) | 23 (37.1%) | 0.922 | 0.337 |
| Gestational age at delivery (weeks) | 39.3 ± 1.0 | 39.2 ± 0.8 | -<br>0.721 | 0.471 |
| Fetal weight (g) | 3101 ± 365 | 3225 ± 353 | 1.067 | 0.287 |
| Birth defects | 1 (1.6%) | 0 (0%) | - | - |
| Insulin use | 2 (3.2%) | 4 (6.5%) | - | 0.680 |

Note: Weight gain during pregnancy was compared using the Mann-Whitney U test; fetal weight and gestational age at delivery were compared using the t-test; cesarean section rate was compared using the chi-square test; insulin use was compared using Fisher's exact test. Statistical
power: 58% for weight gain (detectable $\Delta \geq 3.5\text{kg}$ ); 42% for cesarean rate (detectable $\Delta \geq 20\%$ ) at $\alpha = 0.05$ .

Given the limited sample size (N = 124, 62 pairs), we performed an observed power analysis for the main continuous outcome (gestational weight gain) and the key binary outcome (cesarean section rate). Under the effect sizes detected in this study (Cohen’s d = 0.25 for continuous data; 15% absolute risk difference for binary data), the estimated statistical power was 58% and 42%, respectively. These values are provided solely to illustrate the reduced ability of the current study to detect small-to-moderate effects; they should not be interpreted as confirmatory evidence.

## Discussion

### Interplay of occupational status and pre-pregnancy BMI

Our study identified a significant association between occupational status and pre-pregnancy BMI (P < 0.05), with employed women exhibiting slightly higher pre-pregnancy BMI values compared to their unemployed counterparts. This observation may reflect the complex interplay of socioeconomic, behavioral, and physiological factors. Employment often imposes time constraints, potentially leading to irregular dietary patterns, sedentary behavior, and sleep deprivation—all established risk factors for weight dysregulation. Furthermore, occupational stress may exacerbate metabolic dysfunction through dysregulated hypothalamic-pituitary-adrenal (HPA) axis activity and chronic cortisol elevation [10, 16]. However, these findings represent an association rather than causation; future longitudinal or interventional studies are needed to delineate causal pathways and quantify the contribution of individual factors (e.g., job-related physical activity, dietary choices).

### Heterogeneity in BMI determinants

Notably, the relationship between employment and BMI exhibited substantial interindividual variability. While employed women may face weight-related challenges, factors such as health literacy, access to recreational facilities, occupational physical demands, and socioeconomic support systems likely modulate this association[17–20]. For instance, some employed women maintain healthy weights through structured exercise regimens or meal prepping, whereas unemployment does not universally confer metabolic protection, particularly in settings with limited health resources or financial constraints.

### Contextual interpretation of findings

The generalizability of our results must be contextualized within the study’s setting—Xiamen, a high-income urban region in China with robust female workforce participation. In low-resource settings, employed women may face compounded stressors (e.g., food insecurity, limited healthcare access), whereas in affluent societies, workplace wellness initiatives (e.g., subsidized gym memberships, healthy catering) may mitigate BMI disparities[21,22]. Cross-cultural comparisons are essential to extrapolate these findings.

### Interpretation of null findings on pregnancy outcomes

“Notably, occupational status showed no significant association with gestational weight gain, cesarean delivery, fetal weight, or other pregnancy outcomes after propensity score matching (Table 4).The absence of significant differences in cesarean delivery rates and fetal weight between employed and unemployed women suggests that under standardized GDM management, occupational status may not be a independent determinant for these particular complications. This highlights the effectiveness of current prenatal care protocols in mitigating potential disparities arising from employment stress.[6]. Future interventions should extend this paradigm by embedding pre-pregnancy BMI screening into occupational health assessments, as advocated by the ADA for high-risk populations.”

### Emerging trends and public health implications

Encouragingly, growing health awareness among working women is reshaping lifestyle norms, evidenced by trends such as lunchtime exercise and healthier food delivery options. Workplace interventions targeting stress reduction (e.g., mindfulness programs, flexible scheduling) should be prioritized, given evidence that psychological stress is a modifiable risk factor for metabolic disease [10]. Employer-sponsored wellness programs could integrate these strategies with prenatal nutrition counseling and flexible exercise opportunities, creating comprehensive support systems for women at risk of GDM. Over time, such initiatives may attenuate BMI disparities related to occupational status.”.

### Implications of sample size and statistical power

The propensity score matching process yielded 62 matched pairs (N=124), which constrained our statistical power. Post-hoc analysis confirmed insufficient power (42–58%) to detect clinically relevant differences in key outcomes (e.g., <20% relative change in cesarean risk or <2 kg difference in gestational weight gain). This aligns with previous PSM studies where rigorous matching often trades sample size for covariate balance. Crucially, however: 1. Directional consistency: All point estimates (e.g., cesarean rate: 27.4% vs 37.1%; fetal weight: 3101g vs 3225g) showed trends favoring unemployed women, suggesting possible undetected effects. 2. Clinical relevance threshold: Observed differences fell below thresholds for clinical significance (e.g., <500g for fetal weight; <5% absolute risk difference for cesarean delivery as per obstetric guidelines [23]. 3. Generalizability trade-off: While reduced power limits causal inferences, the high covariate balance (SMD<0.1) enhances internal validity for the employed-unemployed contrast within this cohort.

Future multi-center studies with larger samples are warranted to validate these observations.

### Study limitations

1. Our findings should be interpreted with consideration of the following limitations:
2. Sample constraints: The single-center design and modest sample size may limit external validity.
3. Occupational classification bias: Dichotomizing employment status (employed/unemployed) overlooks nuances such as job type, work intensity, or shift patterns [24]
4. Residual confounding: Despite rigorous PSM implementation [25], residual confounding from unmeasured variables (e.g., dietary habits) may persist, a limitation inherent to observational designs. [26]
5. Cross-sectional design: Causality cannot be inferred; prospective cohorts or randomized trials are needed to establish temporal relationships.
6. The primary limitation remains the modest post-matching sample size, reducing power to detect modest effect sizes. However, this reflects the trade-off between covariate balance (achieved via strict PSM caliper=0.15) and statistical power—a well-documented challenge in matched cohort studies

## Conclusion

In this propensity score-matched analysis, occupational status was associated with pre-pregnancy BMI but not with other pregnancy outcomes (gestational weight gain, cesarean delivery, or fetal weight) in women with GDM. “Our results underscore the necessity of pre-pregnancy BMI control in working women, aligning with the ADA Standards of Care (2025) that prioritize pre-conception weight optimization as a key strategy for reducing GDM complications [3]. The observed association between occupational status and pre-pregnancy BMI suggests that workplace health programs should integrate targeted pre-conception counseling, particularly for employed women at high risk of GDM.” Future research should investigate causal mechanisms and evaluate interventions tailored to occupational contexts. **Clinical relevance**: Routine pre-pregnancy BMI screening and employer-based wellness initiatives are warranted for women at risk of GDM.

## Ethical considerations

This retrospective study used anonymized data and was granted an exemption from formal ethics approval by the Ethics Committee of Xiamen University Affiliated Xiang’an Hospital (Waiver No. XAHLL2025028).

## Data availability statement

TheAll relevant data are within the manuscript and its Supporting Information files.

